# Boston Cognitive Assessment (BOCA) — a comprehensive self-administered smartphone- and computer-based at-home test for longitudinal tracking of cognitive performance

**DOI:** 10.1101/2021.10.08.21264757

**Authors:** Andrey Vyshedskiy, Rebecca Netson, Elisabeth Fridberg, Priyanka Jagadeesan, Matthew Arnold, Sophie Barnett, Anjali Gondalia, Victoria Maslova, Lauren deTorres, Simone Ostrovsky, Danijel Durakovic, Andrei Savchenko, Sienna McNett, Mikhail Kogan, Irene Piryatinsky, Dov Gold

## Abstract

Longitudinal cognitive testing is essential for developing novel preventive interventions for dementia and Alzheimer’s disease; however, the few available tools have significant practice effect and depend on an external evaluator. We developed a self-administered 10-minute at-home test intended for longitudinal cognitive monitoring, Boston Cognitive Assessment or BOCA. The goal of this project was to validate BOCA. BOCA uses randomly selected non-repeating tasks to minimize practice effects. BOCA evaluates eight cognitive domains: 1) Memory/Immediate Recall, 2) Language Comprehension/Prefrontal Synthesis, 3) Visuospatial Reasoning / Mental rotation, 4) Executive function / Clock Test, 5) Attention, 6) Mental math, 7) Orientation, and 8) Memory/Delayed Recall. BOCA was administered to patients with cognitive impairment (n = 50) and age- and education-matched controls (n = 50). Test scores were significantly different between patients and controls (p < 0.001) suggesting good discriminative ability. The Cronbach’s alpha was 0.87 implying good internal consistency. BOCA demonstrated strong correlation with Montreal Cognitive Assessment (MOCA) (R= 0.90, p <0.001). The study revealed strong (R=0.94, p <0.001) test-retest reliability of the total BOCA score one week after participants’ initial administration. The practice effect tested by daily BOCA administration over 10 days was insignificant (β=0.03, p=0.74). BOCA has the potential to reduce the cost and improve the quality of longitudinal cognitive tracking essential for testing novel interventions designed to reduce or reverse cognitive aging. BOCA is available online gratis at www.bocatest.org.

## Introduction

Many treatable health conditions (e.g., sleep disorders, hypertension, diabetes, heart failure, hypothyroid), deficiencies (e.g., vitamin B12), as well as lack of movement and social interactions can affect memory and thinking ^1–3^. Longitudinal monitoring of cognitive health can help clinicians assess if an underlying condition is causing cognitive decline and guide timely therapeutic interventions ^4^. In addition, longitudinal monitoring is essential for testing novel interventions designed to reduce or reverse cognitive aging ^5^. Standard cognitive assessments are not suited for monthly cognitive evaluations. First, they ubiquitously rely on trained professionals. While this approach has a high sensitivity and specificity for the detection of dementia, it is time and resource intensive. Second, the number of variations of standard tests is often limited resulting in practice effects ^6^. Therefore, there is a clear need for a self-administered cognitive test that can be repeated periodically and is resistant to practice effects ^7^. Such a test could be performed at home or in the clinic by using randomly selected non-repeating tasks to minimize practice effects.

In the last two decades, the availability of computerized cognitive testing with diagnostic accuracy comparable to traditional pen-and-paper neuropsychological testing have improved significantly ^8^. The National Institutes of Health Toolbox Cognition Battery ^9^, the Cognitive Stability Index ^10^, CogState ^11^, BrainCheck ^12^, Neurotrack ^13^, CNS Vitals ^14^ can replace existing paper screening tests like Montreal Cognitive Assessment (MOCA), but they still require a trained evaluator administering the test to a patient. At home approaches have also been developed and validated (e.g., the Computer Assessment of Mild Cognitive Impairment ^15^, COGselftest ^16^, and MicroCog ^11^). These tests showed good neuropsychological parameters, but were primarily designed for single use cases and are not validated for longitudinal cognitive tracking ^8^. To the best of our knowledge, to date, only one test has been specifically designed for at-home longitudinal cognitive monitoring. The *Brain on Track* self-administered web-based test for longitudinal cognitive assessment was developed in 2014, in Portugal ^17^. However, the *Brain on Track* was not available in English as of 2021. Therefore, we aimed to develop an online self-administered test for longitudinal cognitive assessment that could be used at home on multiple devices including smartphones, tablets, and computers.

Previously, BOCA has been validated against and shown strong correlations (*r* = 0.80, p < 0.01) with the Telephone Interview for Cognitive Status (TICS) ^18^. BOCA demonstrated good internal consistency (Cronbach’s alpha= 0.79), adequate content validity, and strong (*r*= 0.89, p <0.001) test-retest reliability of the total BOCA score one week after participants’ initial administration. This study aimed to assess the convergent validity of the BOCA with the MOCA test in a larger group of participants, and provide further evidence of the BOCA’s validity and reliability.

## Methods

Boston Cognitive Assessment or BOCA is a 10-minute, self-administered online test that uses randomly selected non-repeating tasks to minimize practice effects. BOCA includes eight subscales: Memory/Immediate Recall, Memory/Delayed Recall, Executive function/Clock Test, Visuospatial Reasoning/Mental rotation, Attention, Mental math, Language/Prefrontal Synthesis, and Orientation, Table 1. The maximum total score is 30, with higher scores indicating better cognitive performance.

**Table 1.**
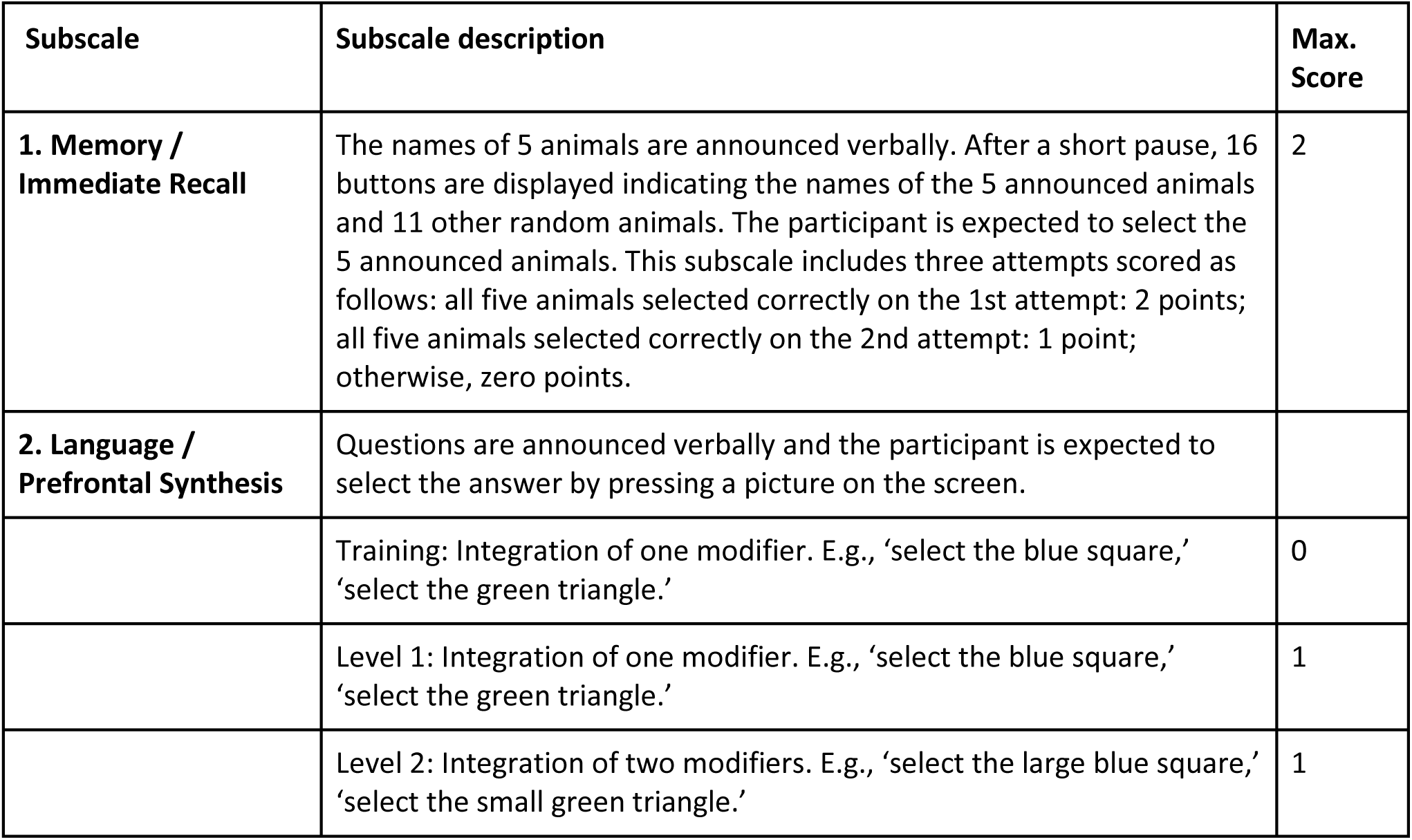
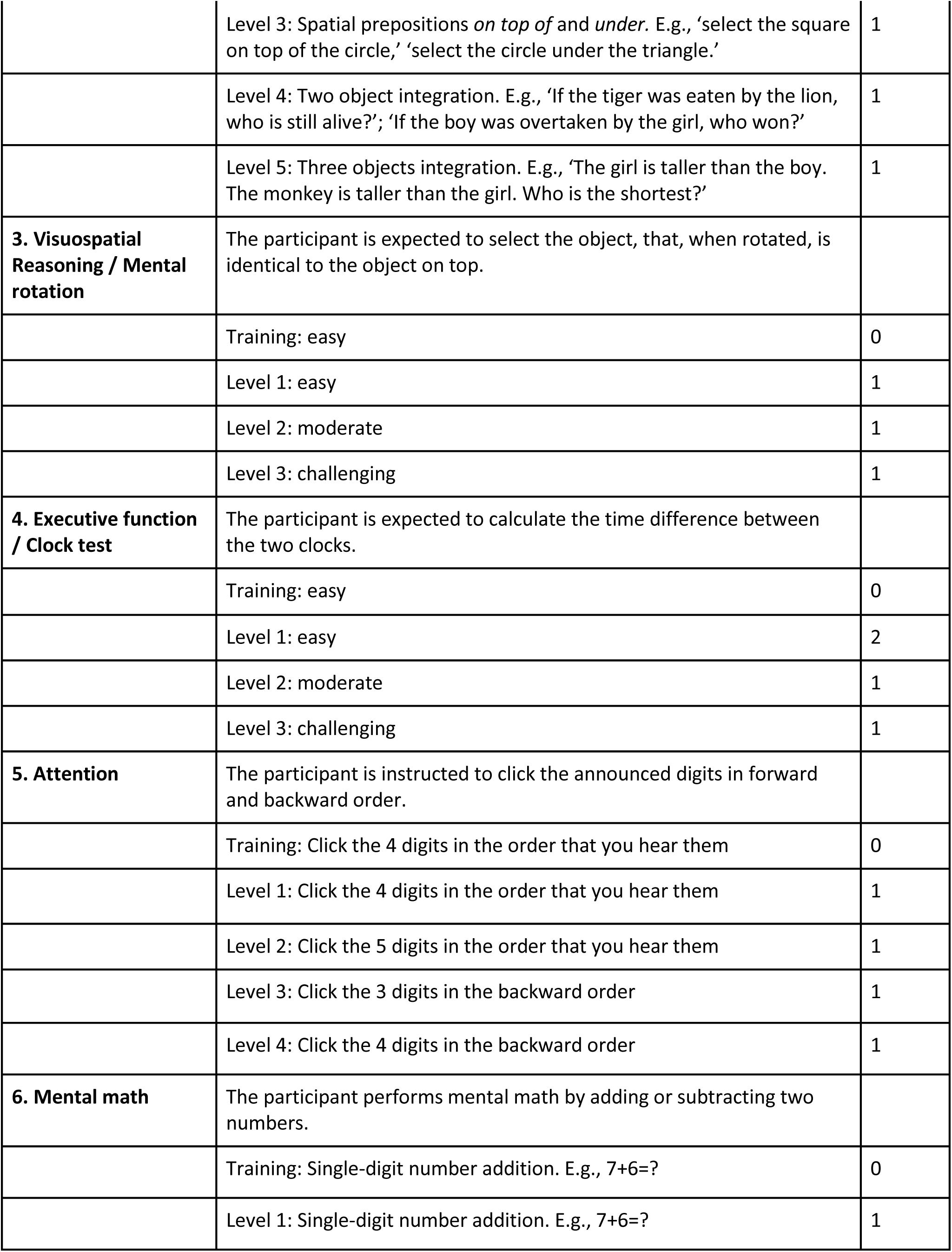
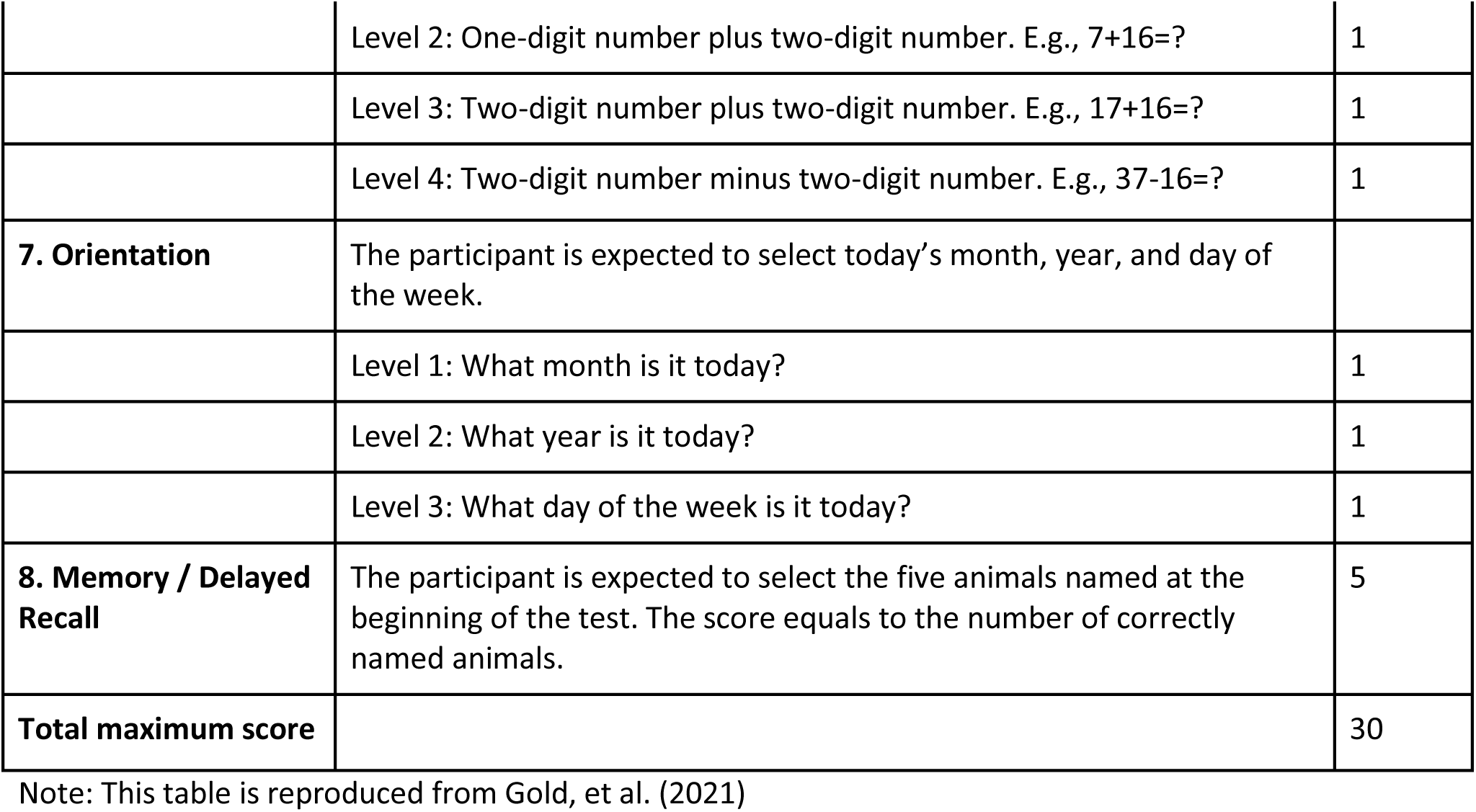
BOCA subscales, example questions, and scoring.

### Principles for test development

As a self-administered test, BOCA was designed to be user-friendly and self-explanatory. The testing within each subscale is preceded by a *training session*, Table 1. The goal of a training session is to familiarize participants with both the task and the answering protocol in each subscale. Training sessions are unscored and are repeated until the participant finds the correct answer. During training sessions, users are provided with the “Correct/Incorrect” feedback. Once a user has confirmed his/her ability to answer a question correctly in a given subscale, they are able to proceed to scored questions of that subscale.

Assessment in each subscale is accomplished by a set of questions with gradually increasing level of difficulty. For example, assessment in the Mental Math subscale has four levels of difficulty: in the first level, the participant is expected to add two single-digit numbers (E.g., 7+6=?); in the second level, one-single-digit number and one two-digit number (E.g., 7+16=?); in the third level, they must add two, two-digit numbers (E.g., 17+16=?); and in the fourth level, they must subtract a two-digit number from another two-digit number (E.g., 37-16=?), Table 1.

Each level begins with a set of announced instructions and has a limited duration of 40 seconds, excluding the tests’ instructions. As BOCA is intended to be used repeatedly, randomly selected non-repeating tasks are used to minimize practice effects.

BOCA was programmed to work on all devices including smartphones and tablets and in all browsers. BOCA is light on data usage transmitted over the web and can be administered over slower 3G connections. The sound check at the beginning of the test ensures that users clearly hear the instructions. Furthermore, users are asked to find a solution in their mind and to avoid using pen and paper.

#### 1. Memory / Immediate Recall

The names of 5 animals are announced verbally. After a short pause, 16 buttons are displayed indicating the names of the 5 announced animals amidst 11 distractor animals. Participants are expected to select the 5 announced animals. This subscale includes three attempts scored as follows: all five animals selected correctly on the 1st attempt: 2 points; all five animals selected correctly on the 2nd attempt: 1 point; otherwise, zero points are given. With 16 answer choices, the probability of selecting five correct animals by chance is 0.02%; four correct animals 1.3%; three correct animals 13%; two correct animals 38%; one correct animal 38%; and zero correct animal 11%.

#### 2. Language / Prefrontal Synthesis

Complex language comprehension and prefrontal synthesis subscale has five levels of difficulty. In level 1, participants are expected to integrate one modifier (colors: red, blue and green) with a noun (geometrical figures: square, triangle, and circle) and click on the corresponding picture on the screen. With 9 answer choices, the probability of selecting the correct answer by chance is 11%.

In level 2, participants are expected to integrate two modifiers (color and size) with a noun (geometrical figures: square, triangle, and circle) and click on the corresponding picture on the screen (E.g., ‘select the large blue square,’ ‘select the small green triangle’). As in level 1, since the three colors, two sizes, and the three geometrical figures are equally familiar, the difficulty is standardized across all possible assessments. With 18 answer choices, the probability of selecting the correct answer by chance is 5.5%.

In level 3, participants are expected to integrate spatial prepositions (*on top of* or *under*) with geometrical figures (square, triangle, and circle) and click on the corresponding picture (E.g., ‘select the square on top of the circle,’ ‘select the circle under the triangle’). In line with previous levels, since the two spatial prepositions and the three geometrical figures are equally familiar, the difficulty is standardized across all possible tasks. With 12 answer choices, the probability of selecting the correct answer by chance is 8.3%.

In level 4, participants are expected to mentally combine two objects as instructed by a semantically-reversible sentence. Sentences in which swapping the subject and the object result in a new meaning are called semantically-reversible sentences. By contrast, in a nonreversible-sentence (e.g., “The boy writes a letter”) swapping the subject and the object results in a sentence with no real meaning (“The letter writes a boy”). With 2 answer choices, the probability of selecting the correct answer by chance is 50%.

In level 5, the participant is expected to mentally combine three objects as instructed by a semantically-reversible sentence (e.g., “The boy is taller than the girl. The monkey is taller than the boy. Who is the shortest?” In this case the girl is the shortest.). With 3 answer choices, the probability of selecting the correct answer by chance is 33.3%.

Correct answers in all levels are scored as one, incorrect answers are scored as zero.

#### 3. Visuospatial Reasoning / Mental rotation

The mental rotation subscale has three difficulty levels. In each level, participants are expected to select an object, that, when rotated, is identical to the object on top. At each level, the task is selected randomly from the pool of 120 shapes. To ensure standardization across assessments, the pool of 120 shapes were carefully selected to be equal in the number of visual features. With 3 answer choices in each difficulty level, the probability of selecting the correct answer by chance is 33.3%. Correct answers in all levels are scored as one, incorrect answers are scored as zero.

#### 4. Executive function / Clock Test

In this subscale, participants are expected to calculate the time difference between two analog clocks. The test has three difficulty levels. In level 1, both minute hands are on the hour and both hour-hands are between 1 and 6 (the difference between hour-hands is no greater than 4 hours).

In level 2, both minute hands are on the hour, whereas one hour-hand is between 1 and 6 and the other is between 7 and 11.

In level 3, one minute-hand is on the hour and the other is on half-hour, whilst one hour-hand is between 1 and 6 and the other is between 7 and 11. Participants respond by separately dialing hours and minutes (the minute-dial uses 5-minutes steps). With 12 hour choices and 12 minute choices on each dial there are 144 answers, the probability of selecting the correct answer by chance is 0.7%.

Correct answers in levels 2 and 3 are scored as one and in level 1 as two, incorrect answers are scored as zero.

#### 5. Attention

Attention testing in BOCA has four levels of difficulty. In level 1, participants are expected to remember and click the four digits in the forward order. The numbers (zero to 9) are randomly drawn from a pre-recorded list. To ensure a consistent difficulty across tasks, mathematical rules were incorporated into the algorithm, for example, the algorithm avoids consecutive numbers 0,1,2,3. With 10 answer choices (zero to 9), the probability of clicking the four digits in the correct order by chance is 0.02%.

In level 2, participants are expected to remember and click five digits in the forward order. With 10 answer choices, the probability of clicking the five digits in the correct order by chance is 0.003%.

In level 3, participants are expected to remember and click three digits in the backward order. With 10 answer choices, the probability of clicking the three digits in the correct order by chance is 0.14%.

In level 4, participants are expected to remember and click four digits in the backward order. With 10 answer choices, the probability of clicking the four digits in the correct order by chance is 0.02%.

Correct answers in all levels are scored as one, incorrect answers are scored as zero.

#### 6. Mental math

Mental math testing in BOCA has four levels of difficulty. In level 1, participants are expected to add two single-digit numbers (E.g., 7+6=?). To ensure standardization across assessments, only numbers between 6 and 9 are used in this task.

In level 2, participants are expected to add one single-digit number and one two-digit number (E.g., 7+16=?). To ensure standardization across assessments, the one-digit number is between 6 and 9 and the two-digit number is between 16 and 19.

In level 3, participants are expected to add two two-digit numbers (E.g., 17+16=?). To ensure standardization across assessments, the first two-digit number is between 16 and 19 and the second two-digit number is between 26 and 29.

In level 4, participants are expected to subtract a two-digit number from another two-digit number (E.g., 37-16=?). To ensure standardization across assessments, the first two-digit number is between 31 and 35 and the second two-digit number is between 16 and 19.

With 21 answer choices (3 rows of 7 digits in each row), the probability of clicking the correct digit by chance is 4.8%. Correct answers in all levels are scored as one, incorrect answers are scored as zero.

#### 7. Orientation

Orientation is tested in BOCA in 3 levels. In level 1, participants are expected to select the current month. With 12 answer choices, the probability of clicking the correct month by chance is 8.3%.

In level 2, participants are expected to select the current year. With 21 answer choices, the probability of clicking the correct year by chance is 4.8%.

In level 3, participants are expected to select the current day of the week. With 7 answer choices, the probability of clicking the correct day of the week by chance is 14.3%.

Correct answers in all levels are scored as one, incorrect answers are scored as zero.

#### 8. Memory / Delayed Recall

In the delayed recall subscale participants are expected to select the five animals named at the beginning of the BOCA test in the subscale 1. The delayed recall score equals to the number of correctly named animals. With 16 answer choices, the probability of selecting 5 correct animals by chance is 0.02%; the probability of selecting 4 correct animals by chance is 1.3%; the probability of selecting 3 correct animals by chance is 13%; the probability of selecting 2 correct animals by chance is 38%; the probability of selecting 1 correct animal by chance is 38%; and the probability of selecting no correct animal by chance is 11%.

### Montreal Cognitive Assessment (MOCA)

BOCA was compared to Montreal Cognitive Assessment (MOCA), a popular pen-and-paper test of global cognition ^19^. The MOCA assesses several cognitive domains: 1) Memory/ Delayed Recall is assessed by learning of five nouns and recalling them 5 minutes later (5 points); 2) Visuospatial abilities are assessed using a number/letter connection exercise (1 point), a three-dimensional cube copy (1 point), and a clock-drawing task (3 points); 3) Multiple aspects of executive functions are assessed using an alternation trail-making task (1 point), a phonemic fluency task (1 point), and a two-item verbal abstraction task (2 points). 3) Naming is assessed using a three-item naming task with animals (lion, camel, rhinoceros; 3 points); 4) Language is assessed using a repetition of two syntactically complex sentences (2 points); 5) Abstract thinking is assessed by asking a patient to describe the similarity between two objects (2 points); 6) Attention is evaluated using a sustained attention task (target detection using tapping; 1 point) and digit repetition forward and backward (1 point each); 7) Arithmetic is assessed by a serial subtraction task (3 points); 8) Temporal and spatial orientation is evaluated by asking the subject for the date and the city in which the test is occurring (6 points). The maximum score is 30 points with higher scores indicating better cognitive performance.

### Participants

Participants were invited to the study online and recruited from the geriatric and memory clinics in eastern Massachusetts and Washington, DC. All of the participants in the study gave their informed consent prior to their inclusion. In participants with cognitive impairment the caregiver consent was also requested.

The overall inclusion criteria for participants was ≥ 25 years of age. Cognitive impairment was defined as the presence of subjective cognitive complaints over a period of at least 6 months reported by the patient or family members and MOCA score of less than 25. No participants were excluded.

### Statistical Analysis

To assess the correlation between BOCA and MOCA test scores, Pearson correlation was utilized with the corresponding p-value determined by the Student’s t-distribution table. The conditions for independent two samples t-test were assessed via the Shapiro-Wilk test for normal distribution. All subtests’ scores met the normality assumption with p > 0.05. The significance level alpha = 0.05 was utilized in concluding if the correlation was significant. Sample estimates are reported as the Pearson correlation coefficient denoted by ‘R’. Confidence Intervals (CI) are indicated at 95%.

Cronbach’s alpha was obtained to determine internal consistency of the BOCA test. It was computed through a two-way mixed effect model, with assumptions that a group of multiple participants is randomly selected from a population. Using this model, the participants and the subscale scores are considered sources of random effects. Subscale scores were determined through pooled scaling. Labels for interpretation of correlational strength were as follows: 0.1–0.3 indicated a small or weak association, 0.31–0.5 indicated a medium or moderate association, and 0.51–1.0 indicated a large or strong association ^20^.

For test-retest analysis, the Pearson correlation was used to measure the strength of the linear relationship between the first BOCA exam and the second BOCA exam. Pearson correlation coefficient is given as the quotient of the standardized form of covariance of the test-retest total scores and the standard deviation of each trial under the normal distribution assumption.

## Results

### Differences in the score between patients and controls

Cognitive impairment was defined as the presence of subjective cognitive complaints over a period of at least 6 months reported by the patient or family members and MOCA score of less than 25. Participants with cognitive impairment (“patients”, n=50) were matched to controls (n=50) based on education obtained and age. Accordingly, there were no significant differences between patients and controls regarding age and education (Table 2).

**Table 2.** Characteristics of participants. The data shown as Mean (SD).

|  | Controls (N=50) | Patients (N=50) | P - Value |
| --- | --- | --- | --- |
| Age | 70.6 (13.1) | 70.0 (14.2) | 0.82 |
| Education (years) | 16.2 (2.3) | 16.1 (2.9) | 0.94 |
| Males | 28% | 36% | 0.14 |

All participants completed the BOCA and MOCA. The average total MOCA score was 26.80 (95% Confidence Interval: 26.26; 27.34) in controls and 18.16 (CI: 16.56; 19.75) in patients. The MOCA total score was statistically different between controls and patients (t (98) = 10.25, p < 0.001), Table 3. All MOCA subscale scores were also significantly different among controls and patients (Table 3).

**Table 3.** MOCA performance in patients and controls.

| MOCA Subscales | Controls | Patients | P-Value |
| --- | --- | --- | --- |
| Visuospatial | 4.33 (0.88) | 2.73 (1.72) | <0.001 |
| Naming | 2.96 (0.20) | 2.67 (0.63) | 0.003 |
| Attention 1 and 2 | 1.96 (0.20) | 1.59 (0.67) | <0.001 |
| Attention 3 | 2.86 (0.41) | 1.78 (1.28) | <0.001 |
| Orientation | 5.84 (0.37) | 4.45 (1.68) | <0.001 |
| Delayed Recall | 3.33 (1.41) | 0.59 (1.04) | <0.001 |
| Language | 2.61 (0.53) | 1.96 (0.94) | <0.001 |
| Abstraction | 1.90 (0.47) | 1.47 (0.79) | 0.001 |
| MOCA Total Score | 26.80 (1.88) | 18.16 (5.63) | <0.001 |

The average total BOCA score was 27.30 (CI: 26.67; 27.94) in controls and 18.28 (CI: 16.35; 20.21) in patients. The BOCA total score was statistically different between controls and patients (t (98) = 8.911; p<0.001). All BOCA subscale scores were also significantly different between the controls and patients, Table 4. There was strong positive statistically significant correlation between the BOCA total score and the MOCA total score with R = 0.90 (CI: 0.86, 0.93), p < 0.001.

**Table 4.**
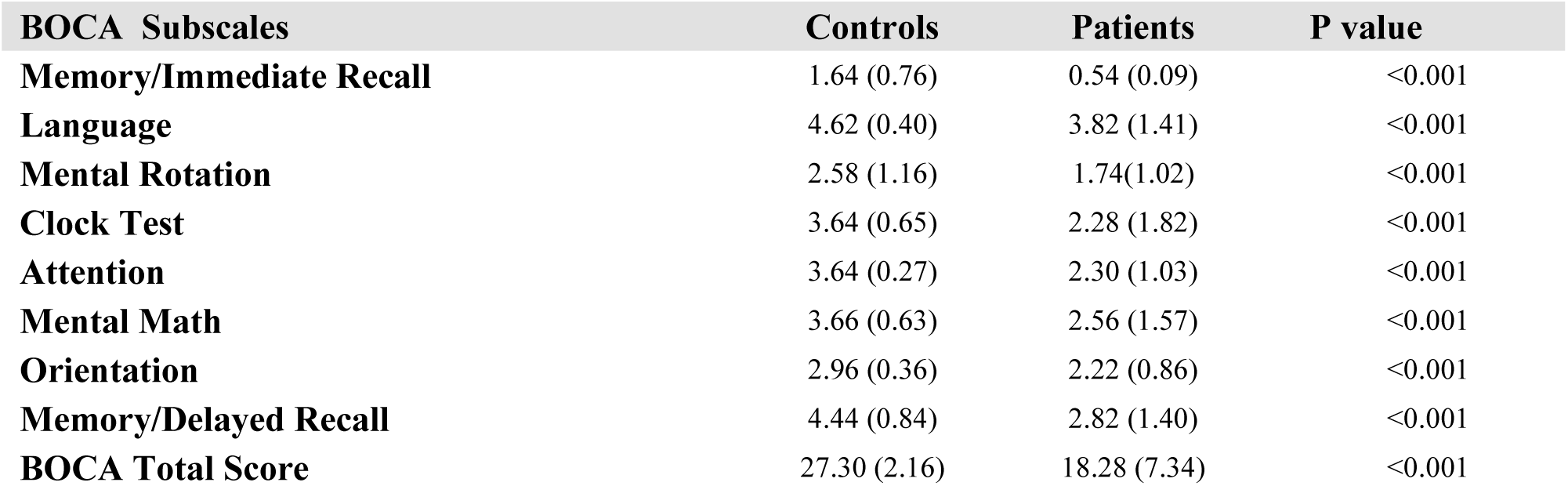
BOCA performance in patients and controls.

Linear regression was computed for patients and controls to assess the relationship between the total BOCA score (independent variable) and their age (dependent variable), Figure 1. As expected, there was a mild negative correlation between the total BOCA score and age in both patients: R = -0.12 (CI: -0.38, 0.15), p = 0.39; and controls: R = -0.29 (CI: -0.52, -0.01), p = 0.04.

**Figure 1:**
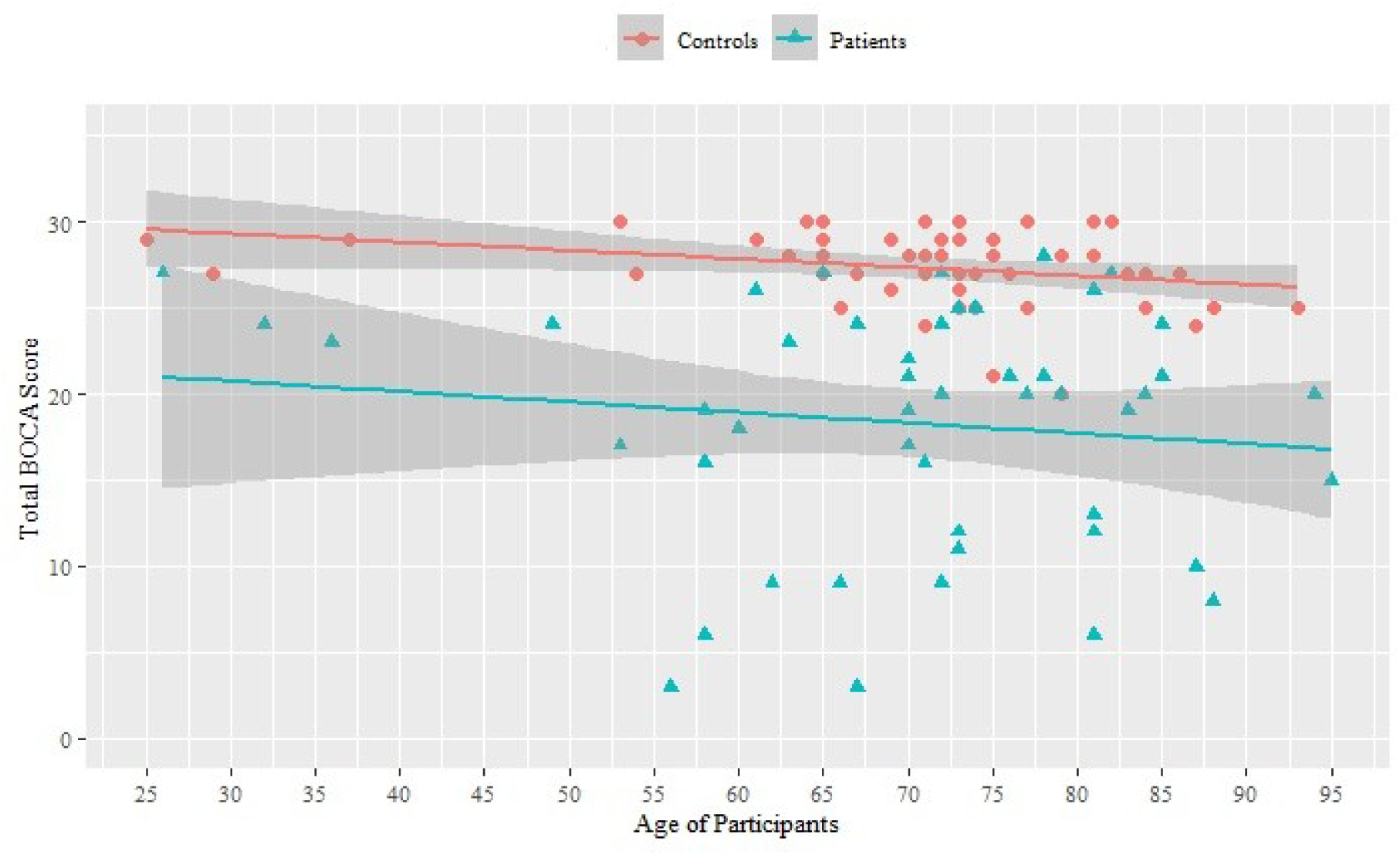
Linear regression between the total BOCA score and age of participants. Confidence bands are shown at 95%.

**Figure 2:**
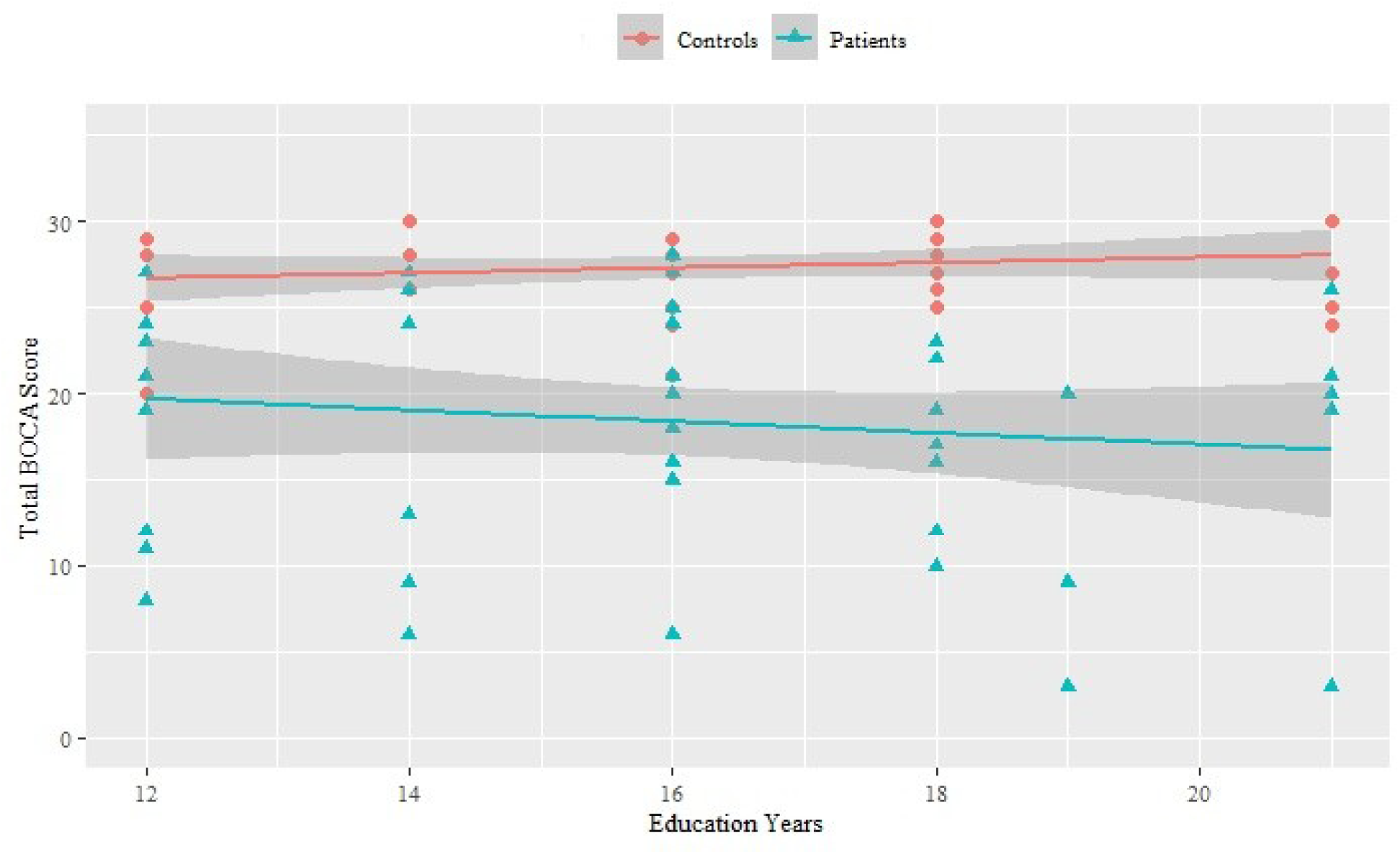
Linear regression between the total BOCA score and education years of participants. Confidence bands are shown at 95%.

Linear regression was also computed for patients and controls to assess the relationship between the total BOCA score and their number of years of education. There was a small insignificant correlation between the total BOCA score and years of education in both patients: R = -0.13 (CI: -0.39, 0.16), p = 0.37; and controls: R = 0.15 (CI: -0.13, 0.41), p = 0.28.

### BOCA Reliability Analysis

The correlation matrix for BOCA subscales was obtained via the Pearson method, Table 5. The strongest correlations were between the Orientation and the Attention subscales (R = 0.62, t(98) = 7.72, p < 0.001),the Orientation and the Clock Test subscales (R = 0.57, t(98) = 6.56; p < 0.001), and the Immediate Memory and the Attention subscales (R = 0.55, t(98) = 6.49; p < 0.001). All but one bivariate correlation was significant at the 0.01 level. The only insignificant correlation at the 0.01 level was between the Delayed Recall and the Language subscales (R = 0.24, t(98) = 2.48, p = 0.015).

**Table 5.** BOCA subscales Pearson correlation matrix. \**p* < 0.05, \*\**p* < 0.01

| BOCA Subscale | Immediate Recall | Language | Mental Rotation | Clock Test | Attention | Mental Math | Orientation | Delayed Recall |
| --- | --- | --- | --- | --- | --- | --- | --- | --- |
| Memory / Immediate Recall | - |  |  |  |  |  |  |  |
| Language | 0.41** | - |  |  |  |  |  |  |
| Mental Rotation | 0.34** | 0.3** | - |  |  |  |  |  |
| Clock Test | 0.42** | 0.45** | 0.36** | - |  |  |  |  |
| Attention | 0.55** | 0.43** | 0.33** | 0.48** | - |  |  |  |
| Mental Math | 0.44** | 0.42** | 0.45** | 0.53** | 0.49** | - |  |  |
| Orientation | 0.50** | 0.44** | 0.41** | 0.57** | 0.62** | 0.49** | - |  |
| Memory / Delayed Recall | 0.52** | 0.24* | 0.39** | 0.49** | 0.50** | 0.35** | 0.53** | - |

Internal consistency of the eight BOCA subscales was assessed using Cronbach’s alpha. Results indicated good internal consistency α = 0.87 (CI: 0.81, 0.90), p < 0.001. Item-total correlation (ITC) was evaluated using Pearson’s product moment coefficient between each subscale score and the total score, Table 6. All subscales demonstrated high (>0.5) ITC. The highest ITC for the Orientation subscale was 0.72 and the lowest ITC for the Mental Rotation subscale was 0.501.

**Table 6.** Factor analysis for BOCA.

| **BOCA Subscales** | **Item-Total Correlation** | **Cronbach's Alpha if Item Deleted** |
| --- | --- | --- |
| **Memory/Immediate Recall** | 0.63 | 0.84 |
| **Language** | 0.51 | 0.85 |
| **Mental Rotation** | 0.50 | 0.85 |
| **Clock Test** | 0.66 | 0.83 |
| **Attention** | 0.68 | 0.83 |
| **Mental Math** | 0.63 | 0.84 |
| **Orientation** | 0.72 | 0.83 |
| **Memory/Delayed Recall** | 0.59 | 0.85 |

One subscale at a time was then removed and the Cronbach’s alpha was re-calculated for the remaining 7 subscales, Table 6. For the purposes of this test, all subscale scores were standardized. The resulting Cronbach’s Alpha were all positive, demonstrated high (>0.83) internal consistency, and remained stable.

The Kaiser-Meyer-Olkin measure of sampling adequacy indicated that the strength of the relationships among subscales was high (KMO = 0.886) thus factor analysis was possible. Factor analysis of the eight BOCA subscales yielded one factor with an eigenvalue of 4.14 accounting for 51.76% of the variance in the data. Factor two had an eigenvalue of 0.801 and accounted for 10.01% of the variance. The remaining six factors had eigenvalues below 0.8. Based on the low amount of variance explained by the second factor, results of the Scree Plot, and moderate to strong positive correlations between all eight BOCA subscales and the first factor, only the first factor was retained. This factor was subsequently identified as global cognitive functioning and encompassed all of the eight subscales.

The BOCA test-retest reliability was evaluated by calculating a Pearson correlation coefficient between the first administration of the BOCA and the re-administration of the BOCA to the same participants approximately one-week later (93 participants), Table 7. The one-week test-retest correlation coefficient for the BOCA total score was R = 0.94 (CI: 0.91 0.96), p < 0.001, revealing excellent BOCA long-term stability. As shown in Table 7, each of the BOCA subscales showed significant test-retest correlations one week after participants initial completion of the test.

**Table 7:** Test-Retest Reliability. \**p* < 0.05, \*\**p* < 0.01

| BOCA Subscales | BOCA 1 | BOCA 2 | Pearson Correlation |
| --- | --- | --- | --- |
| **Memory/Immediate Recall** | 1.11 (0.96) | 1.23 (0.93) | 0.68** |
| **Language** | 4.32 (0.92) | 4.10 (1.12) | 0.50** |
| **Mental Rotation** | 2.04 (1.12) | 2.24 (0.96) | 0.52** |
| **Clock Test** | 3.06 (1.31) | 3.13 (1.32) | 0.58** |
| **Attention** | 2.84 (1.41) | 2.84 (1.44) | 0.82** |
| **Mental Math** | 2.90 (1.40) | 2.92 (1.38) | 0.84** |
| **Orientation** | 2.60 (0.81) | 2.57 (0.91) | 0.53** |
| **Memory/Delayed Recall** | 3.59 (1.66) | 3.71 (1.59) | 0.70** |
| **Total Score** | 22.52 (7.52) | 22.75 (7.74) | 0.94** |

Practice effect was assessed by administering BOCA daily to 10 participants and analyzed through a linear mixed effect model, where the trial number was modeled as a fixed effect, participants as random effects, and test score as the dependent variable. An interaction term was added to account for the effect of each trial nested within each individual participant. The effect of the trial number on the BOCA total score was insignificant (β=0.03, SE=0.08, p=0.74), Figure 3. The fixed effect intercept of the trial number had β = 17.7, SE = 2.5, p<0.0001. The effect of the trial number on individual BOCA subscales was also insignificant, Table 8.

**Figure 3.**
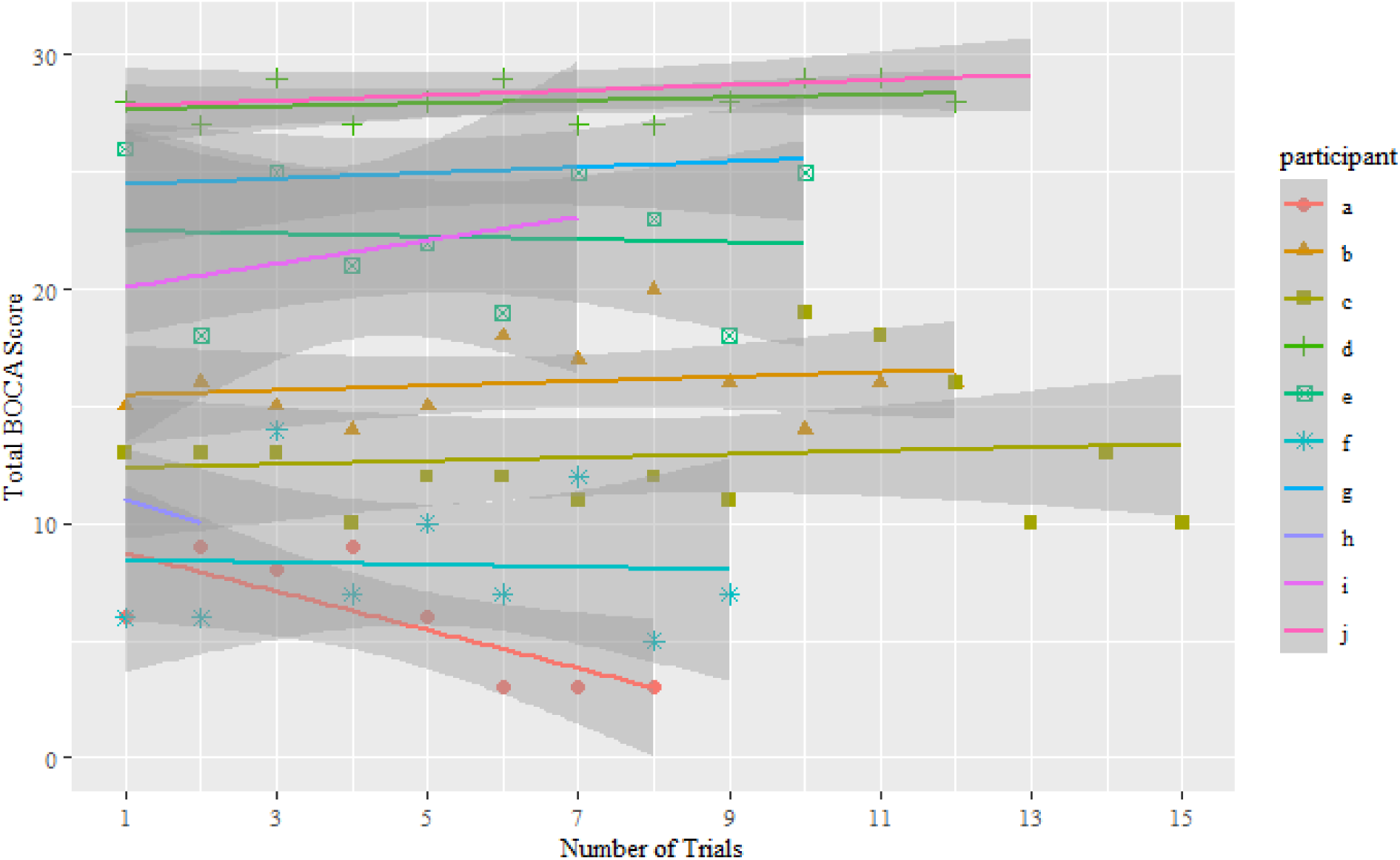
Practice effect of the daily BOCA administration in 10 participants.

**Table 8:** Practice effect analysis.

| BOCA Subscales | Beta ($\beta$) | Standard Error | T Statistic ($\beta/SE$) | P-Value |
| --- | --- | --- | --- | --- |
| Memory/Immediate Recall | 0.01 | 0.02 | 0.64 | 0.52 |
| Language/PFS | -0.03 | 0.04 | -0.68 | 0.52 |
| Mental Rotation | 0.01 | 0.03 | 0.48 | 0.67 |
| Clock Test | 0.01 | 0.02 | 0.64 | 0.53 |
| Attention | 0.02 | 0.02 | 0.74 | 0.48 |
| Mental Math | 0 | 0.03 | -0.15 | 0.89 |
| Orientation | 0.02 | 0.03 | 0.67 | 0.53 |
| Memory/Delayed Recall | 0 | 0.03 | 0.26 | 0.80 |
| Total Score | 0.03 | 0.08 | 0.34 | 0.74 |

## Discussion

This manuscript discusses the implementation of the Boston Cognitive Assessment or BOCA, a self-administered at-home test used for cognitive screening and longitudinal monitoring. The manuscript shows the results of BOCA’s validation procedure in 50 participants with cognitive impairment (patients) and 50 controls matched by education obtained and age. Test scores were significantly different between patients and controls (p < 0.001) suggesting good discriminative ability. Internal consistency of BOCA was high (Cronbach’s alpha = 0.87) and BOCA exhibited excellent test-retest reliability (R =0.94). In further support of the BOCA’s validity, exploratory factor analysis yielded a single factor explaining a plurality of the variance in participants scores. This factor was felt to reflect global cognitive functioning, and suggests the test’s internal structure matches the construct intended to be measured. Finally, practice effect, assessed by administering BOCA daily for 7 days, was insignificant (β=0.03, SE=0.08, p=0.74) confirming the absence of learning even when BOCA is used daily.

These results agree with previously reported observations comparing BOCA to the Telephone Interview for Cognitive Status (TICS) that demonstrated BOCA’s ability to differentiate patients from controls (p<0.001), good BOCA’s internal consistency (Cronbach’s alpha = 0.79), strong correlations with the TICS (r = 0.80, p < 0.01), and strong (r= 0.89, p <0.001) test-retest reliability of the total BOCA score one week after participants’ initial administration ^18^. Furthermore, this study reconfirmed the factor structure of the BOCA in a novel sample, and demonstrated that education does not significantly influence patient scores, which is a common limitation of other global screening instruments. Although not included in this study, previous evaluations of the BOCA found it to possess equal or better sensitivity (79%) and specificity (90%) to mild cognitive impairment relative to other global screening tests ^18^.

BOCA was designed to be a self-administered and self-explanatory test. BOCA experience starts with receiving an email or SMS with a link to the test. Clicking on the link, automatically opens the browser with BOCA introductory page or the BOCA app on an iPhone. The mandatory sound check ensures that patients can hear the instructions. The testing in each subscale is preceded by a training session in order to familiarize participants with both the task and the answering protocol. Training sessions are unscored and are repeated until the participant finds the correct answer. Once a user has confirmed his/her ability to answer correctly in a given subscale, they proceed to scored questions of that subscale. The testing in each subscale is accomplished by a set of questions which gradually increase in difficulty. Randomly selected non-repeating tasks are used to minimize practice effects.

BOCA achieves high sensitivity to cognitive impairment by using eight orthogonal subscales: Memory/Immediate recall, Memory/Delayed recall, Executive function/Click test, Visuospatial reasoning/Mental rotation, Attention, Mental math, Language/Prefrontal synthesis, and Orientation, Table 3. In selecting BOCA subscales, we strived to cover as many neurologically distinct cognitive processes as possible. The variety of cognitive tasks had to be balanced against the test duration. The maximum total score is 30, with higher scores indicating better cognitive performance. The resulting BOCA takes an average 11.0±1.8 minutes to complete and therefore satisfies our preset duration specification.

### BOCA advantages and limitations

The BOCA test shares potential advantages with the other computerized cognitive tests: convenience and cost-effectiveness, reduction of the examiner bias, reduction of performance anxiety, automatic recording and storing of the results, and progress tracking ^8,17^. The main criticisms of at-home cognitive tests relate to the presence of potential technical difficulties faced by older adults ^8^. To address this concern, BOCA uses unscored training sessions in each subscale, large unambiguous buttons, and an internet browser to present the test. No installation of any files is required and the test can be taken on any device, including smartphones and tablets. All participants in this trial have taken the test at home with minimal help from their caretakers, suggesting good usability in the at-home setting. As the number of adults using smartphones increases, the lack of familiarity should decrease as well.

However, unsupervised testing at home also creates the potential issue of patient non-compliance. For example, patients could record names of animals on paper or use a calculator in the math subscale. In the future, it may be possible to add eye tracking and face-movement recognition to avoid this potential issue.

Computerized tests performed at home also have potential technological hurdles, such as different hardware and internet speed of the patients’ computers. Using an internet browser to present the test (instead of locally installed software) guarantees homogeneity in the different platforms and simplifies the procedure for patients. Once patients receive an email or SMS from their provider, they click on the link, automatically opening the browser with the test. Furthermore, BOCA preloads all of the data needed for each subscale before its initiation, resulting in independence from internet speed. A slower 3G network can affect the waiting time for the test to start, but not the duration of the test.

High internal consistency (Cronbach’s alpha = 0.87), excellent test–retest reliability, good discriminative ability, and the absence of practice effect validate BOCA as an effective cognitive test for longitudinal clinical use. BOCA is the first self-administered at-home test intended for cognitive monitoring. It has the potential to revolutionize cognitive tracking essential for testing novel interventions designed to reduce or reverse cognitive aging. Additionally, the test can be used to assess the effect of anesthesia, long-term effect of cancer drugs, COVID fog, and other conditions known to affect cognition. BOCA is available online gratis at www.bocatest.org

## Data Availability

Data are available from the corresponding author upon reasonable request.

## Author affiliations

Boston University, USA: Andrey Vyshedskiy, Elisabeth Fridberg, Priyanka Jagadeesan, Matthew Arnold, Anjali Gondalia

Alzheimer’s Light, Miami, FL, USA: Rebecca Netson, Sophie Barnett, Victoria Maslova, Lauren deTorres, Simone Ostrovsky, Danijel Durakovic, Andrei Savchenko

Department of Clinical Psychology, William James College, Newton, MA, USA: Dov Gold

Center for Integrative Medicine, George Washington University, USA: Sienna McNet, Mikhail Kogan

Neuropsychological Assessment Clinic, Boston, MA, USA : Irene Piryatinsky

## Compliance with Ethical Standards

The research has been approved by the George Washington University Committee on Human Research, USA, Institutional Review Board (IRB). All methods were performed in accordance with the relevant guidelines and regulations.

## Acknowledgments

We wish to thank Dr. C. Prather for insightful comments on this manuscript.

## Author contributions

Study design: AV,MK, IP. Developed the BOCA test: AV. Implementation of the BOCA test: AV, AS, DD, LdT, SO. Acquisition of data: EF, PJ, MA, SB, AG, VM, SM, IP. Analysis of data: RN. Drafting the manuscript : AV. Manuscript revision for important intellectual content: AV, RN, DG, MK, PJ. All authors have read and approved the manuscript.

## Availability of data and materials

Data are available from the corresponding author upon reasonable request.

